# Smoking Cessation Efforts for Patients with Asthma and COPD

**DOI:** 10.64898/2026.02.14.26345148

**Authors:** Shira Yellin, Marcus Rauhut, Eric Kutscher, Edward Anselm

## Abstract

Smoking Cessation Efforts for Patients with Asthma and COPD

**Introduction:** Smoking cessation can alter the natural history of both COPD and asthma by reducing the frequency and severity of exacerbations and slowing disease progression. Accordingly, the Global Initiative for Asthma and the Global Initiative for Chronic Obstructive Lung Disease recommend that clinicians address smoking cessation at every visit using counseling and pharmacotherapy.

**Methods:** The Mount Sinai Health System includes seven hospitals and more than 400 outpatient locations in the New York metropolitan area, all using a unified electronic medical record (Epic). De-identified data from calendar year 2024 were extracted for individuals identified as current smokers via the EMR smoking status tool. Patients with asthma and/or COPD were identified using ICD-10 codes. Tobacco treatment was defined as receipt of counseling or pharmacotherapy, including varenicline, bupropion, or nicotine replacement therapy.

**Results:** Among 961,997 patients, 58,566 (6.1%) were identified as current cigarette smokers. Across all health system encounters, 32.6% of smokers with both asthma and COPD were given any treatment, followed by 26.7% of smokers with COPD, 13.0% of smokers with asthma, and 9.9% of cigarette smokers without these conditions. Smokers seen in pulmonary clinics were the most likely to be given treatment (17.4%), followed next by primary care (6.6%).The most commonly used treatment for all cohorts and all treatment settings was nicotine with the exception of the pulmonary clinic where varenicline predominated.

**Discussion:** Despite higher treatment rates among smokers with asthma and COPD, only one-third of those with either condition received cessation treatment over a full year, underscoring the need for sustained system-wide quality improvement efforts.

## Introduction

Cigarette smoking is well known to exacerbate asthma and Chronic Obstructive Pulmonary Disease (COPD), resulting in increased emergency room visits and hospitalizations.^1-4^ The Global Initiative for Chronic Obstructive Lung Disease (GOLD) identifies cigarette smoking as both the most easily modifiable risk factor for COPD, and recommends encouragement of cessation for all individuals who smoke.^5^ To make this operational, Mirza et al explicitly call for the treatment of tobacco use disorder at every visit.^6^

The National Asthma Education and Prevention Program (NAEPP) *Key Clinical Activities for Quality Asthma Care* states “No patient with asthma should smoke or be exposed to Environmental Tobacco Smoke (ETS).”^7^ Physicians should review smoking status at the initial visit and all subsequent visits and, if patients smoke or are regularly exposed to ETS, should encourage and refer them to stop smoking.”^8^ The Global Initiative for Asthma (GINA) recommends addressing all modifiable risk factors, which includes smoking, as part of the routine cycle of assessment-treatment-review in asthma care.^9^

Clinical practice guidelines updated in 2008 outline evidence-based treatment that can be provided at every outpatient visit, regardless of patients’ readiness to change.^10^ This guideline also states, “Counseling and medication are effective when used by themselves for treating tobacco dependence. The combination of counseling and medication, however, is more effective than either alone.”^10^ The World Health Organization released a guideline for smoking cessation in adults in 2024, which recommends “brief advice (between 30 seconds and 3 minutes per encounter) be consistently provided by health-care providers as a routine practice to all tobacco users accessing any health-care settings.”^11^

Treatment options for tobacco cessation include counseling and medications. Counseling can be divided by the amount of time spent by the clinician: 0-3 minutes (no available CPT code), 3-10 minutes (CPT 99406) or greater than ten minutes (CPT 99497). CMS allows reimbursement for up to eight sessions of counseling per year for CPT codes 99406 or 99407. Most health insurance plans follow this reimbursement model.^12^ Counseling is also available from a patient’s health plan or from the New York State Quitline. Several medications have been approved for the treatment of tobacco, including nicotine replacement, varenicline, and bupropion, with varenicline shown to be the most efficacious of the three.^13,14^ There is also data to support the addition of a second medications if treatment with one medication is not successful.^15^ All medications are available without cost-share for those with commercial health insurance or Medicaid. Medicare and Medicare Advantage plans cover all prescription medications, but do not cover nicotine patches or the 2mg version of nicotine gum, as these products are available over the counter. The NYS Quitline provides nicotine patches for patients who meet their criteria.

Despite these guidelines and effective medications for smoking cessation, performance of US clinicians in treating tobacco use is poor,^16^ with frequent recommendations to adopt systems change approaches.^17,18^ The most recent National Health Interview Survey stated that two out of three cigarette smokers want to quit, yet among those who have seen a doctor in the last year, only about half were provided assistance in smoking cessation.^19^ Of those who made a quit attempt, only 36.3% used an FDA approved medication to quit and only 7.3% received counseling, with an overall success of 8.8%.^19^

Two recent abstracts, based on data from Mount Sinai Health System (MSHS) Electronic Medical Record (EMR), show that patients with asthma receive treatment at a higher rate than patients without this condition, but that there may be many missed opportunities for treatment within both populations.^20,21^

Treatment of people who smoke cigarettes, especially for those with underlying respiratory disease is an important measure of clinical care and health system performance. The National Commission for Quality Assurance (NCQA) is implementing new HEDIS measures for Tobacco Use Screening and Cassation Intervention in 2026 and this analysis can serve as a useful baseline.^22^

## Methods

MSHS consists of seven hospitals and over 400 treatment locations in the New York Metropolitan area. Medical encounters are captured in a single EMR (Epic) De-identified data was extracted for adults over 18, with one or more ambulatory visit, identified as currently smoking either through EMR Smoking Status tool or with an ICD-10 of current smoking with a any treatment visit within the System for the calendar year 2024, including inpatient, outpatient, telemedicine, and emergency department contexts. Patients with asthma, COPD, or both were identified using ICD-10 codes.

Treatment of tobacco documented in the EMR included medications and counseling. For medications, any prescription for varenicline and any prescription for a nicotine replacement therapy (NRT) were considered evidence of treatment with NRT reported in aggregate. Given the multiple indications for bupropion, only prescriptions of bupropion sustained release 150mg twice daily (the approved dosing for smoking cessation) was considered evidence of treatment. For smoking cessation counseling, CPT4 counseling codes 99406 and 99407 were considered evidence of treatment.

Treatment was further evaluated through the creation of an indicator of treatment intensity. Each patient received a score from zero to three: one point for a single medication, one point for a second medication, and one point for counseling. The sum of the scores was divided by the number of patients in the population to obtain the intensity. Treatment intensity was calculated for the groups with asthma only, COPD only, both asthma and COPD, and those without respiratory disease. Intensity was also calculated for different specialty clinics in which tobacco use is commonly treated.

Descriptive statistics for percentages of the population receiving any of the treatment scenarios were computed. Due to the nonnormal distribution of the data, a nonparametric test, specifically the Kruskal-Wallis test, was used. The level of significance was set at P□<□.05, and all tests were 2-tailed. The Kruskal-Wallis test identified global differences between the diagnosis groups and clinic types, followed by Dunn’s post-hoc pairwise comparison. A chi-square test of independence was used to evaluate whether diagnoses were associated with treatment types. Patients with either asthma, COPD or both were compared to other smokers. Two-sided Pearson chi-square p-values were adjusted for multiple comparisons using the Bonferroni correction. We used SPSS version 28 (IBM) for all analyses.

This cross-sectional study was exempt from institutional review board approval given the de-identified nature of the data. The data was assembled by the Internal Data Handling Committee which is part of the Mount Sinai Enterprise Data Services organization. Committee membership includes representatives from the Epic Research Reporting Group, consisting of Physician Informatics, Data Scientists, Data Engineers, and Data Analysts. This multidisciplinary structure ensures appropriate oversight of data preparation, de identification, and release.

## Results

The total population of adults with at least one ambulatory encounter in 2024 was 961,997, with 58,566 (6.09%) identified as smokers. Within this cohort identified as smokers, 4,467 individuals had COPD (7.62%), 6,532 had asthma 11.2, and 2,389 had both asthma and COPD (4.08%).

The demographics of the population of all smokers, smokers with asthma only, smokers with COPD only, smokers with both COPD and asthma and smokers with neither asthma nor COPD (other smokers) is described in Table 1. Notably, patients with COPD were significantly older than the general population of smokers, while the patients with asthma only appear to be similar to the general population. Half (49.6%) of smokers reported their race as “unknown/declined” or “other,” and 43% identified their ethnicity as “unknown/declined.”

**Table 1.** Demographics of patients identified as cigarette smokers in 2024.

|  |  | All Smokers | Asthma and | Asthma only | COPD only | Other Smokers |
| --- | --- | --- | --- | --- | --- | --- |

|  |  |  | **COPD** |  |  |  |
| --- | --- | --- | --- | --- | --- | --- |
| Age | 18-29 | 7,276 (12.4%) | 6 (0.3%) | 1,120 (17.1%) | 18 (0.4%) | 6,132 (13.6%) |
|  | 30-44 | 14,468 (24.7%) | 91 (3.8%) | 1,995 (30.5%) | 104 (2.3%) | 12,278 (27.2%) |
|  | 45-64 | 22,743 (38.8%) | 1,201 (50.3%) | 2,565 (39.3%) | 1,643 (36.8%) | 17,334 (38.4%) |
|  | 65+ | 14,079 (24.0%) | 1,091 (45.7%) | 852 (13.0%) | 2,702 (60.5%) | 9,434 (20.9%) |
| Ethnicity | Hispanic | 8,356 (14.3%) | 608 (25.4%) | 1,480 (22.7%) | 573 (12.8%) | 5,695 (12.6%) |
|  | Not Hispanic or Latino | 25,032 (42.7%) | 1,280 (53.6%) | 2,827 (43.3%) | 2,329 (52.1%) | 18,596 (41.2%) |
|  | Unknown/Patient Declined | 25,178 (43.0%) | 501 (21.0%) | 2,225 (34.1%) | 1,565 (35.0%) | 20,887 (46.2%) |
| Race | American Indian or Alaska Native | 79 (0.1%) | 2 (0.1%) | 13 (0.2%) | 3 (0.1%) | 61 (0.1%) |
|  | Asian | 2,676 (4.6%) | 50 (2.1%) | 230 (3.5%) | 92 (2.1%) | 2,304 (5.1%) |
|  | Black or African-American | 9,521 (16.3%) | 758 (31.7%) | 1,622 (24.8%) | 803 (18.0%) | 6,338 (14.0%) |
|  | Native Hawaiian or Pacific Islander | 76 (0.1%) | 0 (0.0%) | 17 (0.3%) | 5 (0.1%) | 54 (0.1%) |
|  | Other | 10,349 (17.7%) | 646 (27.0%) | 1,627 (24.9%) | 764 (17.1%) | 7,312 (16.2%) |
|  | Unknown/Patient Declined | 18,693 (31.9%) | 303 (12.7%) | 1,511 (23.1%) | 1,115 (25.0%) | 15,764 (34.9%) |
|  | White | 17,172 (29.3%) | 630 (26.4%) | 1,512 (23.1%) | 1,685 (37.7%) | 13,345 (29.5%) |
| Sex | Female | 25,179 (43.0%) | 1,398 (58.5%) | 3,597 (55.1%) | 1,900 (42.5%) | 18,284 (40.5%) |
|  | Male | 33,360 (57.0%) | 991 (41.5%) | 2,932 (44.9%) | 2,567 (57.5%) | 26,870 (59.5%) |
|  | Unknown | 27 (0.0%) | 0 (0.0%) | 3 (0.0%) | 0 (0.0%) | 24 (0.1%) |
| Total |  | 58,566 (100%) | 2,389 (100%) | 6,532 (100%) | 4,467 (100%) | 45,178 (100%) |

Utilized treatment for smoking cessation outlined by patient diagnosis groups is described in Table 2. Across all encounters in all settings, 32.6% of smokers with asthma and COPD were given any treatment, followed by 26.7% of smokers with COPD and 13.0% of smokers with asthma, each significantly greater than the 9.9% of smokers without any of these respiratory diseases (p<0.05).

**Table 2.** Opportunities for treatment taken: Inclusive of inpatient, emergency room, and clinic settings.

|  | **All Smokers** | **Asthma and COPD** | **Asthma only** | **COPD only** | **Other Smokers** |
| --- | --- | --- | --- | --- | --- |
| Any Treatment | 7,306 (12.5%) | 780 (32.6%)* | 852 (13.0%)* | 1,194 (26.7%)* | 4,480 (9.9%) |
| **Medication Only** | 6,203 (10.6%) | 668 (28.0%)* | 738 (11.3%)* | 982 (22.0%)* | 3,815 (8.4%) |
| Nicotine Only | 3,279 (5.6%) | 350 (14.7%)* | 393 (6.0%)* | 542 (12.1%)* | 1,994 (4.4%) |
| Varenicline Only | 786 (1.3%) | 120 (5.0%)* | 91 (1.4%)* | 169 (3.8%)* | 406 (0.9%) |
| Bupropion Only | 1,465 (2.5%) | 56 (2.3%) | 177 (2.7%) | 124 (2.8%) | 1108 (2.5%) |
| Nicotine and Varenicline Only | 368 (0.6%) | 89 (3.7%)* | 41 (0.6%)* | 95 (2.1%)* | 143 (0.3%) |
| Nicotine and Bupropion Only | 223 (0.4%) | 31 (1.3%)* | 28 (0.4%) | 32 (0.7%)* | 132 (0.3%) |
| **Counseling Only** | 718 (1.2%) | 58 (2.4%)* | 79 (1.2%) | 121 (2.7%)* | 460 (1.0%) |
| **Medication and Counseling** | 385 (0.7%) | 54 (2.3%)* | 35 (0.5%) | 91 (2.0%)* | 205 (0.5%) |
| No Treatment | 51,260 (87.5%) | 1,609 (67.4%)* | 5,680 (87.0%)* | 3,273 (73.3%)* | 40,698 (90.1%) |
| Total | 58,566 (100%) | 2,389 (100%) | 6,532 (100%) | 4,467 (100%) | 45,178 (100%) |

\* p < 0.05 Pearson chi-square tests comparing diagnosis to Other Smokers (p values adjusted using Bonferroni correction)

Among the treatment options, nicotine alone was the most common across all categories 14.7% of smokers with asthma and COPD were given any treatment, followed by 12.0% of smokers with COPD and 6.00% of smokers with asthma, each significantly greater than the 4.4% of smokers without any of these respiratory diseases. The number of nicotine prescriptions was more than three times the varenicline prescriptions across all groups. Optimal treatment, with both medication and counseling, was not employed frequently with the exception of the COPD only group (2.8%) and those patients with both asthma and COPD (3.5%).

Further stratification by medical specialty for each encounter is outlined in Table 3. Overall, smokers received some form treatment during only 3.3% of their outpatient encounters. Patients in the pulmonary clinics received significantly greater intensity of services than all other settings, and were most commonly prescribed varenicline in this setting, compared to nicotine in all other clinic types. Internal Medicine and Primary Care clinics also were significantly better than average (p<0.05). The average intensity of service provided in Cardiology and Psychiatry clinics was significantly better than the other medical and surgical clinics (p<0.05). Notably the lowest rate of treatment was in Allergy clinic.

**Table 3.**
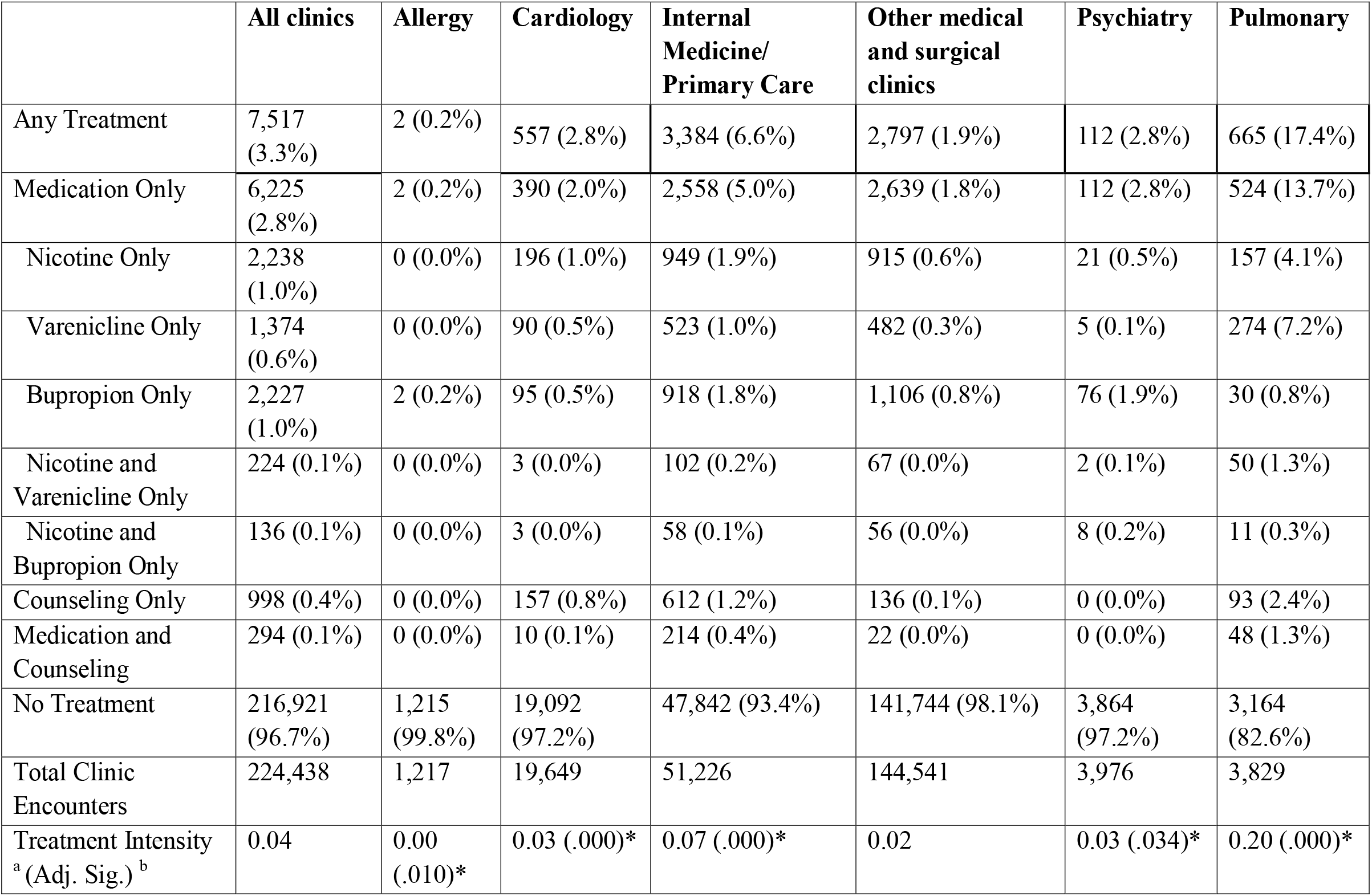
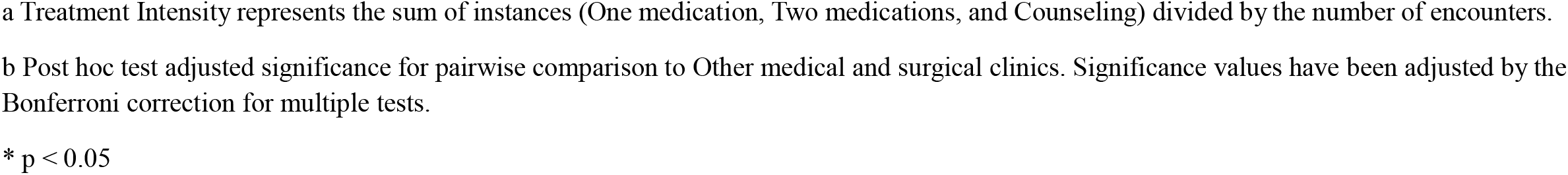
Treatment opportunities taken in clinics by clinic type based on encounters.

|  | **All clinics** | **Allergy** | **Cardiology** | **Internal**<br>**Medicine/**<br>**Primary Care** | **Other medical**<br>**and surgical**<br>**clinics** | **Psychiatry** | **Pulmonary** |
| --- | --- | --- | --- | --- | --- | --- | --- |
| Any Treatment | 7,517<br>(3.3%) | 2 (0.2%) | 557 (2.8%) | 3,384 (6.6%) | 2,797 (1.9%) | 112 (2.8%) | 665 (17.4%) |
| Medication Only | 6,225<br>(2.8%) | 2 (0.2%) | 390 (2.0%) | 2,558 (5.0%) | 2,639 (1.8%) | 112 (2.8%) | 524 (13.7%) |
| Nicotine Only | 2,238<br>(1.0%) | 0 (0.0%) | 196 (1.0%) | 949 (1.9%) | 915 (0.6%) | 21 (0.5%) | 157 (4.1%) |
| Varenicline Only | 1,374<br>(0.6%) | 0 (0.0%) | 90 (0.5%) | 523 (1.0%) | 482 (0.3%) | 5 (0.1%) | 274 (7.2%) |
| Bupropion Only | 2,227<br>(1.0%) | 2 (0.2%) | 95 (0.5%) | 918 (1.8%) | 1,106 (0.8%) | 76 (1.9%) | 30 (0.8%) |
| Nicotine and<br>Varenicline Only | 224 (0.1%) | 0 (0.0%) | 3 (0.0%) | 102 (0.2%) | 67 (0.0%) | 2 (0.1%) | 50 (1.3%) |
| Nicotine and<br>Bupropion Only | 136 (0.1%) | 0 (0.0%) | 3 (0.0%) | 58 (0.1%) | 56 (0.0%) | 8 (0.2%) | 11 (0.3%) |
| Counseling Only | 998 (0.4%) | 0 (0.0%) | 157 (0.8%) | 612 (1.2%) | 136 (0.1%) | 0 (0.0%) | 93 (2.4%) |
| Medication and<br>Counseling | 294 (0.1%) | 0 (0.0%) | 10 (0.1%) | 214 (0.4%) | 22 (0.0%) | 0 (0.0%) | 48 (1.3%) |
| No Treatment | 216,921<br>(96.7%) | 1,215<br>(99.8%) | 19,092<br>(97.2%) | 47,842 (93.4%) | 141,744 (98.1%) | 3,864<br>(97.2%) | 3,164<br>(82.6%) |
| Total Clinic<br>Encounters | 224,438 | 1,217 | 19,649 | 51,226 | 144,541 | 3,976 | 3,829 |
| Treatment Intensity<br><sup>a</sup> (Adj. Sig.) <sup>b</sup> | 0.04 | 0.00<br>(.010)* | 0.03 (.000)* | 0.07 (.000)* | 0.02 | 0.03 (.034)* | 0.20 (.000)* |

a Treatment Intensity represents the sum of instances (One medication, Two medications, and Counseling) divided by the number of encounters.
b Post hoc test adjusted significance for pairwise comparison to Other medical and surgical clinics. Significance values have been adjusted by the Bonferroni correction for multiple tests.
\* $p < 0.05$

Table 4 shows the treatment for patients only with respiratory disease in the various clinic settings. Treatment intensity was somewhat higher in all settings for patients with these diagnoses and was significantly higher than the group without these diagnoses in pulmonary and general medicine clinics.

**Table 4.** Treatment intensity for patients with Asthma, COPD or both showing opportunities taken by clinic type based on encounters.

|  | **All clinics** | **Allergy** | **Cardiology** | **Internal Medicine/**<br>**Primary Care** | **Other medical and surgical clinics** | **Psychiatry** | **Pulmonary** |
| --- | --- | --- | --- | --- | --- | --- | --- |
| Any Treatment | 2,221 (4.8%) | 0 (0.0%) | 138 (3.3%) | 873 (8.1%) | 690 (2.5%) | 37 (3.3%) | 483 (18.6%) |
| Medication Only | 1,824 (3.9%) | 0 (0.0%) | 93 (2.2%) | 650 (6.0%) | 648 (2.4%) | 37 (3.3%) | 396 (15.2%) |
| Nicotine Only | 697 (1.5%) | 0 (0.0%) | 48 (1.2%) | 289 (2.7%) | 237 (0.9%) | 8 (0.7%) | 115 (4.4%) |
| Varenicline Only | 606 (1.3%) | 0 (0.0%) | 25 (0.6%) | 162 (1.5%) | 205 (0.8%) | 3 (0.3%) | 211 (8.1%) |
| Bupropion Only | 363 (0.8%) | 0 (0.0%) | 18 (0.4%) | 142 (1.3%) | 157 (0.6%) | 23 (2.1%) | 23 (0.9%) |
| Nicotine and Varenicline Only | 101 (0.2%) | 0 (0.0%) | 1 (0.0%) | 33 (0.3%) | 30 (0.1%) | 2 (0.2%) | 35 (1.3%) |
| Nicotine and Bupropion Only | 47 (0.1%) | 0 (0.0%) | 0 (0.0%) | 21 (0.2%) | 15 (0.1%) | 1 (0.1%) | 10 (0.4%) |
| Counseling Only | 300 (0.6%) | 0 (0.0%) | 42 (1.0%) | 171 (1.6%) | 32 (0.1%) | 0 (0.0%) | 55 (2.1%) |
| Medication and Counseling | 97 (0.2%) | 0 (0.0%) | 3 (0.1%) | 52 (0.5%) | 10 (0.0%) | 0 (0.0%) | 32 (1.2%) |
| No Treatment | 44,051 (95.2%) | 436 (100.0%) | 4,009 (96.7%) | 9,940 (91.9%) | 26,483 (97.5%) | 1,069 (96.7%) | 2,114 (81.4%) |
| Total Clinic Encounters | 46,272 | 436 | 4,147 | 10,813 | 27,173 | 1,106 | 2,597 |
| Treatment Intensity <sup>a</sup><br>(Adj. Sig.) <sup>b</sup> | 0.05 | 0.00 (.210) | 0.03 (.431) | 0.09 (.000)* | 0.03 | 0.04 (1.00) | 0.22 (.000)* |

<sup>a</sup> Treatment Intensity represents the sum of instances (One medication, Two medications, and Counseling) divided by the number of encounters.
<sup>b</sup> Adjusted significance for pairwise comparison to Other medical and surgical clinics. Significance values have been adjusted by the Bonferroni correction for multiple tests.
\* $p < 0.05$

In all settings, the rate of counseling or counseling with medication is lower than the use of medication alone by a factor of four or greater (Tables 2,3,4).

## Discussion

The calculated prevalence of tobacco use in the ambulatory population at MSHS, 6.09%, is significantly lower than the prevalence of smoking in Manhattan as reported by the New York State Department of Health (NYSDOH) for 2021 (6.09 vs 9.8).^23^ Other counties served by MSHS have even higher prevalence with Brooklyn at 10.7%, the Bronx at 11.4%, Queens at 9.3% and Richmond at 14.7%. The prevalence of smoking in the NYS Medicaid population is reported at 19.1%.^23^ Medical claims and EMRs are well known to under-report the prevalence of tobacco use.^24,25^ Under-reporting of tobacco use status might predispose to over-reporting of intervention rates. Another dimension of under-reporting is the inability of most EMRs to support the capture of each of the many forms of nicotine such as cigars, pouches, or electronic cigarettes, nor do they capture dual use. According to the NYSDOH, an additional 2.7% of New Yorkers use e-cigarettes daily.^23^ Regular screening of patients in ambulatory settings is a key step in effective treatment of tobacco.^26,27^ The exemplar for data capture is Kaiser Permanente of Northern California, which has published their approach and results.^27^

Overall treatment rates of people who smoke cigarettes at the MSHS (12.5%) are consistent with prior research using data from the EMR showing vast undertreatment. Bailey et al extracted data from 143 safety net clinics using Epic. Over the three-year study period, 79% of the patients had their tobacco use status assessed. Of the patients identified as smokers, 46.2% received no treatment, 35% received counseling only, 7.5% received medication only, and 11.1% received both medication and counseling.^28^ Similar patterns emerge from reports based on medical claims: Geletko et al reported on three years data from 2015-2018 National Ambulatory Medical Care Survey, in which 22.1% of smokers received medication or counseling^29^. White looked at data from the 2019 Medical Expenditure Panel and found that 17% received a medication or counseling for the treatment of tobacco.^30^

Undertreatment of tobacco use is repeatedly demonstrated through patient surveys. The most recent National Health Interview Survey reported that among cigarette smokers who have seen a doctor in the last year, only about half were provided assistance in smoking cessation.^19^ Of those who made a quit attempt, only 36.3% used an FDA approved medication to quit and only 7.3% received counseling.^19^ The NCQA Consumer Assessment of Health Plan Satisfaction (CAHPS) consistently show higher rates of treatment,^31^ and results of NCQA 2023 evaluation of Medicaid plans show that 73.5% of tobacco users received advice to quit, 46.6% recall discussion of strategies to quit, and 52.8% recall discussion of a medication to quit.

There are significant methodological differences among these studies, patient surveys and the EMR data. Malloy et al looked at prevalence of tobacco use in the New York State Medicaid program from a number of measurement perspectives and found that using medical claims produced the lowest prevalence and the highest rates of treatment.^24^ Physicians in outpatient settings do not enter ICD-10 codes for nicotine dependence unless they are treating it during that visit. Another explanation for our lower rates may be missed opportunities to bill, document, and/or diagnose tobacco use disorder counseling. Brief counseling may have occurred, or counseling that exceeded three minutes was not billed with CPT code 99406. The MSHS reported a 0.8% rate of use of this code overall and a 2.1% rate in primary care, consistent with data from other academic medical centers.^25^

The MSHS data consistently shows higher rates of treatment for COPD, asthma, and COPD/asthma than the general smoking population. Bailey et al reported the odds ratio of receiving both medication and counseling as higher for COPD and asthma (1.7) than any other condition evaluated.^28^ Tibuakuu et al. looked at Medical Expenditure Panel Survey data from 2006 to 2015 and reported that patients who smoke and have COPD, were 2.86 times more like to receive advice to quit and 1.88 times more likely receive medications.^32^ In a twelve year study of patients with asthma, smoking cessation was as recommended during 21.7% of visits in which the patient was an active smoker.^33^

The gap between recommendations to treat tobacco on every visit and actual performance has been investigated extensively. The barriers identified include clinician lack of knowledge, attitude, skill, self-efficacy, perception of time constraints, smoking cessation not considered a priority, lack of support system, lack of accountability at the level of clinician, practice, or health insurance as well as low reimbursement and lack of economic incentive. A review of barriers to treatment of tobacco patients with COPD recapitulates many of the points raised in the general population.^34^ The literature on effective solutions has been collated by the CDC Office on Smoking and Health,^35^ but implementation has been limited.^36,37^ One promising approach is through the identification and development of institutional champions to leverage systems transformation.^38^ There is also strong evidence that modifications to electronic medical records can enhance the frequency and quality of tobacco treatment.^39,40^

The variation in performance of the medical clinics as shown on table 4 is not surprising. Almaaitah et al., demonstrated significant variation in outcomes of tobacco treatment using medical records to identify 955,420 smokers: among practice sites, adjusted quit rates ranged from 4.5% to 66.4%.^41^ Pulmonologists and general practice physicians, who spend the most time treating patients with asthma and COPD, have the best adoption of treatment practices. ^42^. Interestingly, allergy/immunology, a field which also treats asthma, did not provide treatment to any of the smokers seen in its clinic.

The spectrum of medications used to treat tobacco is large and includes over the counter and prescription nicotine products, bupropion and varenicline. Coverage without copayment for all medications is required under the Affordable Care Act, however Medicare and Medicare Advantage Plans do not cover nicotine patches or gum because of their availability over the counter. In our study, nicotine prescribing (any formulation) exceeded varenicline in every setting except for pulmonary clinics. Bailey et al reported on 2022 data showing that almost a quarter of patients identified as using tobacco had at least one order for a cessation medication, with nicotine replacement therapy being most common (16%), followed by bupropion (8.2%) and varenicline (4.9%).^43^ The most recent National Health Interview Survey reports that among the tobacco users able to quit, 36% used a medication, with the most commonly used medication being a nicotine patch (19.6%), followed by nicotine gum or lozenge (18.4%), varenicline (9.6%), bupropion (6.4%), and nicotine spray or inhaler (1.0%).^44^

Varenicline is the preferred primary treatment for tobacco use disorder.^13,45^ In a study of national pharmacy dispensing trends sponsored by Pfizer in 2018, Chantix, the brand name for varenicline, was prescribed 74.5% of the time a medication was prescribed for the treatment of tobacco use for patients with commercial insurance, 81.8% with Medicare and 40.7% with Medicaid coverage.^46^ Although varenicline is well established as superior to other types of medication, use of this medication has been diminished by as much as 50% after Pfizer removed the medication from the marketplace during September 2021 and generic versions of varenicline became available.^47^ Kahn et al have reported on recent trends in varenicline use and state that varenicline use dropped after the voluntary recall of Chantix by Pfizer in mid-2021 and has not returned to pre-recall levels.^48^ A report on use of varenicline prescribing from a commercial database shows that the drop in varenicline has continued into mid-2023, with prescribing for varenicline now equal in frequency to that of nicotine products.^48^

Although optimal treatment would involve both counseling and medication, this did not occur with any significant frequency. The MSHS data show that medication and counseling were documented on 0.7% of encounters. Several recent reports suggest that academic medical centers are not billing for tobacco treatment even when services are being provided^20,49^, and that the aggregate value of cessation services if provided at every visit would be substantial^49,50^. For example, EMR data from WellSpan Health, a system in Pennsylvania and Maryland, showed that if every eligible tobacco cessation encounter were billed, the three-year reimbursement potential would total $5.9 million.^50^

The impact of smoking cessation for value-based contracts may be even greater as it is one of the few preventive services that is both cost-effective and produces a direct return on investment ^51,52^. A 2024 Wisconsin study showed that a systems change approach to smoking cessation, including the hire of tobacco treatment specialists, showed that patients in a smoking cessation registry had an net reduction of $35 in monthly health care costs^53^ for patients listed in a smoking registry. Using a prevalence rate of 10% the impact on a health system would be $3.50 PMPM (per member per month). Additional savings are likely in subsequent years.^54^

Treatment of tobacco use is at an important threshold. Although the number of people who use cigarettes continues to decline, a wide variety tobacco products of unknown safety are increasingly available.^19^ With the elimination of the CDC Office on Smoking and Health and related cuts to public health programs, important programs that educate citizens, support treatment, and measure progress in cessation have been terminated.^55^ This places added burdens on clinicians and health systems. The introduction of standardized measures for the screening and treatment of tobacco by NCQA^22^ is an important step forward but can only be successful with more detailed studies of clinician performance using EMR data.

### Limitations

Data collected is limited to 2024, and smoking status may have changed during that period. Smoking status is not consistently captured, which would inflate treatment rates. The data on medication is based on prescriptions written and does not indicate whether the prescription was filled. The report did not differentiate among types of nicotine products prescribed. There is no indication of treatment outcomes. Counseling may occur at greater frequency than reported. Patients may have been referred to the New York State Quitline, but this type of data is not captured.

## Conclusions

The MSHS performance in the treatment of tobacco is consistent with national reports. Poor performance in provision of tobacco cessation services is demonstrated by a variety of sources such as the National Health Interview Survey,^44^ medical claims data,^24^ and papers based on electronic medical records.^43,50^ These studies also identify wide variation in performance of individual clinicians, medical clinics, and medical groups.^41,56^ Nationally, we are quite far from the standards of clinical practice guidelines which recommend that patients who smoke should receive treatment at every visit, regardless of a patient’s readiness to change.^10^

Although patients with asthma and or COPD receive more intensive treatment, numerous opportunities to provide tobacco treatment services were missed. This leads to worsening diseases states for patients, as well as missed opportunities for reimbursement for the healthcare system.

Proper documentation of smoking status in the EMR is the first step in ensuring all smokers are identified for tobacco cessation interventions. EMR data can also provide useful and actionable insights into the performance of a health system’s treatment of tobacco.

## Data Availability

All data produced in the present study are available upon reasonable request to the authors

